# Low levels of protective humoral immunity following mild or asymptomatic infection with SARS-CoV-2 in a community-based serological study

**DOI:** 10.1101/2021.03.19.21253982

**Authors:** Thomas W. McDade, Amelia Sancilio, Richard D’Aquila, Brian Mustanski, Lauren A. Vaught, Nina L. Reiser, Matthew P. Velez, Ryan R. Hsieh, Daniel T. Ryan, Rana Saber, Elizabeth M. McNally, Alexis R. Demonbreun

**Author notes:** Corresponding author: Thomas McDade, PhD, Northwestern University, 1810 Hinman Avenue, Evanston, IL 60208, Phone: 847/467-4304.

## Abstract

The degree of protective humoral immunity after mild or asymptomatic SARS-CoV-2 infection is not known. We measured antibody-mediated neutralization of spike protein-ACE2 receptor binding—a surrogate measure of protection against SARS-CoV-2 infection—in a large and diverse community-based seroprevalence study. Comparisons were made across three groups of seropositive participants that differed in the severity of infection and engagement with clinical care (N=790). The clinical group was seropositive for prior infection, symptomatic, and diagnosed with COVID-19 by a healthcare provider. The symptomatic group was seropositive and reported one or more symptoms of infection but received no clinical care. The asymptomatic group was seropositive but reported no symptoms. 86.2% of all infections were mild or asymptomatic; 13.8% received clinical care. Of the clinical cases, 96.3% were outpatient; only 3.7% required hospitalization. Moderate or high levels of neutralizing activity were detected following 27.5% of clinical infections, in comparison with 5.4% of symptomatic and 1.5% of asymptomatic infections. The majority of infections in the general population are mild or asymptomatic and likely result in low levels of antibody-mediated protective immunity.

## Introduction

Between March 15, 2020, and March 10, 2021, COVID-19 was the cause of 528,829 confirmed deaths in the US (1). While severe cases of COVID-19 have strained the healthcare system, the majority of infections with SARS-CoV-2—the virus that causes COVID-19—result in mild or asymptomatic cases that do not lead to hospitalization (2, 3). The level of immune protection resulting from mild or asymptomatic cases is not known. Most studies have focused on immune responses to more severe hospitalized COVID-19 infection, or responses in people with higher or repeated occupational exposure to SARS-CoV-2 (e.g., healthcare workers). Ascertaining the level of protective immunity in the general population is important for determining progress toward herd immunity, as well understanding durability of protection from re-infection.

The objective of this report is to document the level of protective immunity in a community-based sample of adults previously infected with SARS-CoV-2. Neutralizing antibodies play an important role in protection because they are long-lasting, and because they can bind to viral proteins and inhibit entry into host cells (4). For SARS-CoV-2, the surface spike protein engages the human angiotensin-converting enzyme 2 (ACE2) receptor to enter host cells, and anti-spike neutralizing antibodies can block this interaction and prevent infection (5, 6). Application of this important marker of protective immunity in community settings has been limited by logistical and technical challenges associated with live virus methods and venous blood collection. We overcome these challenges by combining in-home collection of finger stick dried blood spot (DBS) samples with a surrogate virus neutralization protocol for quantifying antibody-mediated inhibition of spike-ACE2 interaction in DBS (7). We document high rates of asymptomatic and mild infection with relatively low levels of neutralizing activity in a community-based cohort of individuals previously exposed to SARS-CoV-2.

## Results

A web-based, “no contact” research platform was implemented to recruit a large community-based sample of adults across the city of Chicago between June 24 and November 11, 2020 (3, 8). Participants reported whether they had experienced any of the following symptoms potentially indicative of COVID-19 infection after March 1, 2020: fever or chills; cough; shortness of breath; headache; muscle or body aches; fatigue or excessive sleepiness; diarrhea, nausea, or vomiting; loss of sense of smell or taste.

Analyses focus on the 17.8% of participants that tested seropositive for prior infection based on the presence of IgG antibodies against the receptor binding domain of SARS-CoV-2 (3, 9).

The sample includes women and men across race/ethnic identities between the ages of 18 and 81 (Table 1). Only 64 of 790 participants were essential health care workers. Comparisons were made across three groups, based on self-reported severity of infection and engagement with clinical care. The “clinical” group included individuals who reported experiencing symptoms of infection, who received care from a healthcare provider (phone/online or in a clinical setting), and who received a diagnosis of COVID-19. The “symptomatic” group reported experiencing one or more symptoms of COVID-19, but did not seek clinical care. The “asymptomatic” group reported no symptoms and did not seek clinical care.

**Table 1.** Descriptive statistics for the three study groups.

| Variable | Total | Asymptomatic | Symptomatic | Clinical |
| --- | --- | --- | --- | --- |
| N | 790 | 314 | 367 | 109 |
| Age, years (SD) | 38.6 (13.2) | 37.8 (13.0) | 38.0 (13.0) | 43.0 (13.9) |
| Female | 55.6 | 51.0 | 57.8 | 57.8 |
| Race/ethnicity |  |  |  |  |
| White | 44.2 | 47.8 | 41.7 | 42.2 |
| Hispanic/Latinx | 21.3 | 13.4 | 24.8 | 32.1 |
| Asian | 23.5 | 29.0 | 23.2 | 9.2 |
| Black | 8.5 | 8.0 | 7.4 | 13.8 |
| Other | 2.5 | 1.9 | 3.0 | 2.8 |
| Healthcare worker | 8.1 | 7.3 | 9.0 | 7.3 |
| Pre-existing condition | 25.7 | 20.7 | 26.7 | 36.7 |
| Smoker | 8 | 8.0 | 7.1 | 11.0 |
| Positive COVID-19 test | 3.9 | 0.0 | 0.3 | 27.5 |
| Number of symptoms (IQR) | 1 (0, 3) | 0 (0, 0) | 2 (1, 4) | 5 (3, 7) |
| % neutralization (IQR) | 8.0 (0, 17.9) | 4.8 (0, 13.6) | 8.0 (0.4, 16.9) | 29.8 (12.7, 53.7) |

Notes: The groups differed significantly in mean age (one-way analysis of variance, $F=7.1$ , $p<0.001$ ), race/ethnicity (Pearson $X^2=37.5$ , $p<0.001$ ), presence of a pre-existing condition ( $X^2=11.2$ , $p<0.01$ ), positive diagnostic COVID-19 test ( $X^2=186.8$ , $p<0.001$ ), median number of symptoms (non-parametric K-sample $X^2=458.9$ , $p<0.001$ ), and median % neutralization (non-parametric K-test $X^2=68.1$ , $p<0.001$ ).

Symptomatic infections, without clinical treatment or diagnosis, were most frequent at 46.5% of all infections. 39.8% of infections were asymptomatic. Only 13.8% of infections were symptomatic with clinical care, and of these only 3.7% required hospitalization (0.5% of all infections). The clinical group reported a median of five symptoms of infection, in comparison with two for the symptomatic group. Median % neutralization of spike-ACE2 binding was significantly higher in the clinical group than the symptomatic group (Wilcoxon rank-sum z=8.54, p<0.001) (Figure 1A). Although the difference was relatively small, neutralization was significantly lower in the asymptomatic than the symptomatic group (z=3.90, p<0.001). In the clinical known COVID-19 group, 27.5% of individuals had moderate or high neutralizing activity following infection (Figure 1B). Only 5.4% of symptomatic and 1.5% of asymptomatic cases show evidence of neutralization at this level. Further, neutralizing activity was indistinguishable from zero for 79.8% of symptomatic and 89.8% of asymptomatic individuals, in comparison with 36.7% of the clinical group.

**Figure 1.**
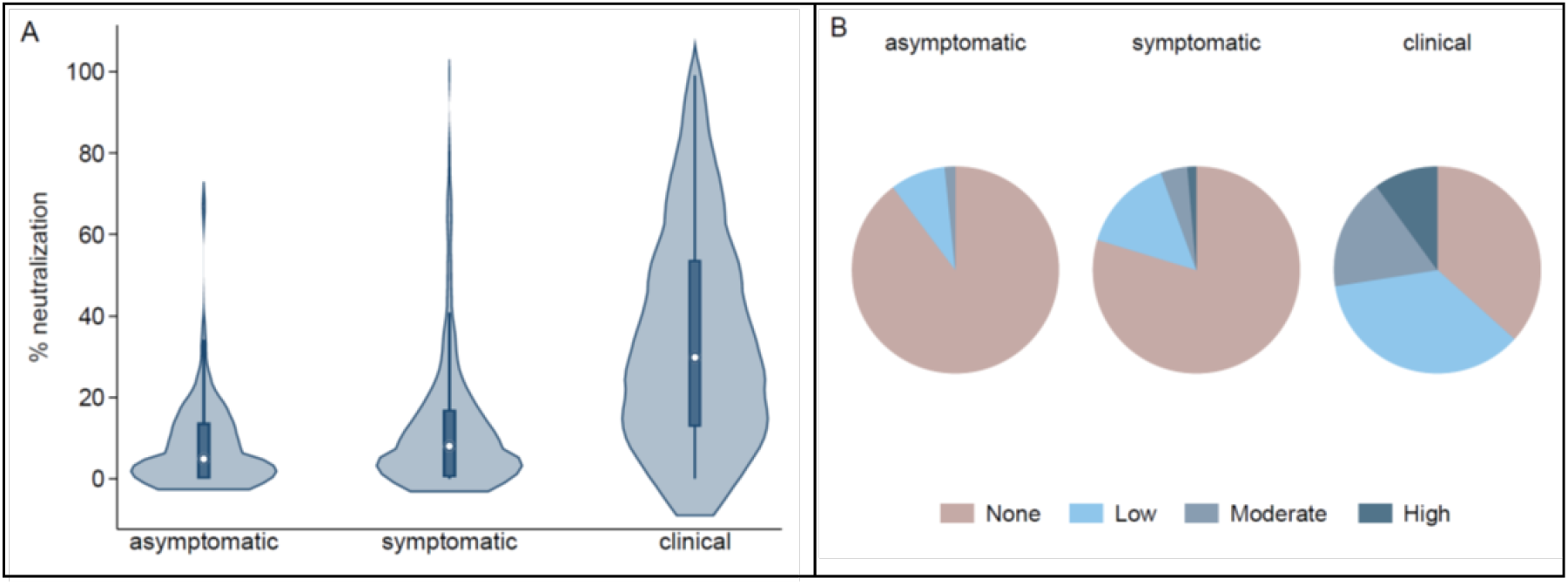
Antibody-mediated neutralization of spike-ACE2 interaction by severity of SARS-CoV-2 infection. A) Violin plot shows median % neutralization and interquartile range, with kernel density, for each group. B) Proportion of cases with low (20-49%), moderate (50-74%), and high (75% and higher) levels of neutralizing activity. The category “none” was defined as % neutralization <20, based on prior analyses of seronegative samples (7).

## Discussion

Two recent studies—one focusing on healthcare workers, the other using clinical records—have reported that prior SARS-CoV-2 infection, as indicated by a positive antibody test, confers protection against future re-infection (10, 11). Similarly, two studies have reported robust responses to the first dose of currently available mRNA COVID-19 vaccines among healthcare workers who tested seropositive prior to vaccination, leading to the proposition that a second vaccine dose may not be needed for previously infected individuals (12, 13).

Our findings suggest caution in applying results from studies of severe cases of COVID-19 and healthcare workers to the general population. We provide evidence for low levels of protective immunity in a large and diverse community-based sample of adults with prior exposure to SARS-CoV-2. Individuals with clinically diagnosed cases of COVID-19 show some evidence of protective immunity, consistent with prior reports. However, these cases comprise less than one in seven infections. The vast majority of infections were mild or asymptomatic, did not result in contact with the healthcare system, and, in this sample, did not generate detectable levels of neutralizing activity in more than 80% of cases. Through 2020, one third of Americans have likely been infected with SARS-CoV-2 (14), but our results suggest that most of these infections did not produce substantial levels of antibody-mediated immunity.

## Materials and Methods

### Participants and study design

Between June 24 and November 11, 2020, N=4,562 adults in the Chicagoland area provided informed consent online, completed a web-based survey, and returned a self-collected finger stick DBS sample in the mail.

Participants retrospectively reported on COVID-19 diagnoses and symptoms experience after March 1, 2020. A pre-existing condition for severe COVID-19 was defined as the presence of chronic obstructive pulmonary disease, diabetes mellitus, cardiovascular disease, or obesity (body mass index >30 kg/m^2^ (15). Smoker was defined by the use of inhaled tobacco products. Participants also indicated sex (based on assignment at birth), primary racial/ethnic identity, and whether they were essential workers in health care. All research activities were implemented under protocols approved by the institutional review board at Northwestern University (#STU00212457 and #STU00212472).

### Surrogate virus neutralizing assay

The competitive immunoassay to quantify neutralizing activity (% neutralization) of spike-ACE2 interaction was previously described (7). DBS samples were available for 790 of 820 seropositive participants, and were incubated with SARS-CoV-2 spike protein and ACE2 conjugated with an electrochemiluminescent label. Neutralizing antibodies, if present, inhibited binding between ACE2 and spike protein, and the Meso Scale Discovery QuickPlex SQ 120 Imager was used to read mean fluorescence intensity (MFI). Percent neutralization was calculated as follows: % neutralization = 100 x 1 - (sample MFI/negative control MFI).

## Data Availability

Data available upon reasonable request from the corresponding author to protect participant confidentiality.

